# Aviation noise, cumulative annoyance and neighborhood ratings: Effect modification of the annoyance dose-response curve from the Neighborhood Environmental Survey

**DOI:** 10.1101/2025.08.02.25332879

**Authors:** Edmund Seto, Ching-Hsuan Huang

## Abstract

The Federal Aviation Administration (FAA) conducted the Neighborhood Environmental Survey (NES) among residents living near 20 airports in the United States (US). This survey aimed to assess exposure to aircraft noise and to update the dose-response curve relating noise exposure to community annoyance. Over 10,000 respondents answered questions related to both noise and non-noise annoyances and provided an overall rating of their neighborhood. Day-night average sound level (DNL) noise exposures for the residential addresses of survey participants were estimated using aviation noise modeling. The NES dose-response curve for high annoyance from aviation noise indicated that more people are likely to be highly annoyed for a given aviation DNL exposure than was estimated from previous dose-response relationships. This paper reports findings from additional analyses of the NES to assess the relationships among noise exposure, cumulative annoyance, and self-reported neighborhood rating. Annoyance from non-aviation noise sources was not found to be a statistically significant modifier of the aviation high annoyance dose-response curve. However, non-noise annoyance was found to modify the dose-response curve, suggesting that individuals cumulatively annoyed may be more sensitive to high annoyance from aviation noise exposure. Principal Components Analysis (PCA) revealed two main components: one loading on all types of noise and non-noise annoyances, and the other loading primarily on aircraft noise annoyance. Neighborhood rating was associated with cumulative annoyance, but less so with aviation noise exposure or aviation noise-related annoyance. These findings suggest the need for further research on cumulative annoyance and the potential synergistic impacts of combined noise exposure from multiple sources on population health.

## Introduction

The relationship between aviation noise exposure and community annoyance is well established. Despite clear evidence of the negative impact of aircraft noise, experts have noted how noise regulation can be a balancing act of protecting public health from the adverse effects of aircraft noise while also protecting aircraft operators and airport agencies from the negative reactions to noise from affected communities – all while keeping transportation services operating for the benefit of society (1). Around the time of the US Noise Control Act of 1972, efforts to understand this balancing act centered on community noise ratings that would indicate the likelihood of a range of community reactions, including varying levels of annoyance, complaints, and legal actions (2). Plots of curves that related day-night average sound levels (DNL or Ldn) to percentages of community complaints evolved into those that estimated the percentages of highly annoyed by DNL noise levels. A significant shift to the use of annoyance measures occurred with the US Federal Interagency Committee on Noise (FICON) declaring in 1992 that annoyance would be the preferred summary measure of adverse community reaction (3).

While noise complaints remain highly relevant and are logged by individual airport noise offices in the US (4), current estimation models of noise annoyance are largely based on dose-response curves (i.e., exposure-response functions). Perhaps the best-known of these is the “Schultz curve”, developed by combining of data from aircraft, road, and rail noise surveys to derive a quadratic function modeling high annoyance in relation to the DNL (5). The original Schultz (1978) study described challenges of noise annoyance curve-fitting, including inter-study variations in the dose-response relationships and conflicting evidence regarding the effect of background noise on individuals’ annoyance response for a given level of, for example, aircraft noise. Some research suggested noise is less disturbing with a high background noise level (i.e., a masking effect), while other studies suggested greater annoyance in areas with higher background levels (i.e., a sensitized community). The effect of combined noise on annoyance remains an unanswered question, with insufficient recent studies providing clear evidence for either masking or sensitization (6). However, it is generally understood that acoustic and non-acoustic factors are important, as are personal, social and environmental contexts.

Since the development of the original Schultz curve and an updated dose-response curve based on a logistic formula in the FICON 1992 report – both of which incorporated various transportation noise sources – efforts have increasingly focused on developing aviation-specific noise annoyance dose-response curves. Notably, Miedema and Oudshoorn fit a separate dose-response relationship for aircraft annoyance, distinguising it from dose-response equations for road traffic and rail (7). However, recent evidence suggests that annoyance may be underestimated by these previous relationships (8–10). With this background, the Federal Aviation Administration (FAA) embarked on new research to update the annoyance dose-response curve for the US.

The FAA’s Neighborhood Environmental Survey (NES) consisted of a mail survey followed by phone interviews of adult residents living around 20 US airports (10). Over 10,000 people responded to the survey, which included questions about annoyance from aircraft noise as well as other noise and non-noise annoyances. The survey also asked people to provide an overall neighborhood rating on an 11-point scale from worst to best. Survey responses were linked to modeled DNL noise estimates. Findings from the FAA’s analysis included an updated dose-response curve for the percentage of high annoyed by aircraft noise based on the aviation DNL noise metric, which estimated higher annoyance levels than previous dose-response curves. Their analysis, however, did not extend to examining the other responses to annoyance responses or the neighborhood rating. Furthermore, their analysis did not focus on the potential shifts in the dose-response curve due to annoyance from other noise sources or non-noise factors.

The objectives of our study were twofold. First, we aimed to confirm the dose-response curve for high-annoyance from aircraft noise using pooled public-use data from the NES. Secondarily, within this objective, we sought to determine if the dose-response is modified by annoyance from other non-aviation noise sources or by overall cumulative annoyance from neighborhood factors. Our second main objective was to assess the relationship between aviation noise and overall neighborhood rating, as well as the relationship between cumulative annoyance and neighborhood rating. We discuss the implications of these analyses in terms of population health impacts on airport communities and future research needs.

## Methods

Our analyses use data collected by the FAA’s NES, which was designed to survey annoyance from aircraft noise in the US. The full methods for the survey are described in the FAA’s technical report (10). Briefly, the NES was carried out on a representative sample of 20 airports selected from a sampling set of 95 US airports that met multiple conditions: (1) location within the contiguous US, (2) at least 100 average daily jet operations, (3) at least 100 people with noise exposure ≥65 dB DNL, and (4) at least 100 people with noise exposure between 60 and 65 dB DNL. Among the airports that met these conditions, three large international airports – Hartsfield-Jackson Atlanta International Airport (ATL), Chicago O’Hare International Airport (ORD), and Los Angeles International Airport (LAX) – were included. Additionally, at least one of the three major New York City airports had to be chosen. The selection also aimed to balance multiple factors, such as maintaining the same proportion of airports as the population within FAA regions, daily temperature, percentage of nighttime flight operations, average daily flight operations, aircraft fleet ratio of commuter to large jet aircraft, and population within 5 miles of the airport. Ultimately, the 20 airports selected were: Albuquerque (ABQ), Albany (ALB), Hartsfield-Jackson Atlanta (ATL), Austin-Bergstrom (AUS), Bradley (BDL), Boeing Field/King County International (BFI), Billings Logan (BIL), Des Moines (DSM), Detroit Metropolitan Wayne County (DTW), McCarran Las Vegas (LAS), Los Angeles (LAX), LaGuardia (LGA), Bill and Hillary Clinton Airport/Adams Field (LIT), Memphis (MEM), Miami (MIA), Chicago O’Hare (ORD), Savannah/Hilton Head (SAV), Norman Mineta San Jose (SJC), Syracuse Hancock (SYR), and Tucson (TUS). Questionnaires were mailed to an initial sample of 26,700 addresses with the goal of obtaining 100 survey respondents in noise exposure strata of 50-55, 55-60, 60-65, 65-70, and 70+ dB DNL at each airport, for a total of 500 responses per airport. Household addresses were sampled from the US Postal Service Computerized Delivery Sequence File within each stratified exposure area. Of the delivered questionnaires (25,018), 40% were completed (10,007). Telephone interviews were also conducted as part of the NES; however, these data were not analyzed for this study. Protocols for the survey were reviewed and approved by the Westat Institutional Review Board. Survey protocols and questionnaire are available in the technical report (10).

Noise exposures for survey respondents’ household addresses were estimated for the NES using the FAA’s Integrated Noise Model (INM) 7.0d and annual flight tracking data specific to each of the selected airports. The DNL noise metric was selected for consistency with previous dose-response curves, such as those developed by the FICON, the International Commission on the Biological Effects of Noise (ICBEN), and the original Schultz curve (3,5,11).

The DNL is also the established metric within the Code of Federal Regulations (CFR), Part 150, for evaluating compatible land use for aircraft noise. Calculated noise exposure was provided for each survey response in five categories: DNL ≤55, 55-60, 60-65, 65-70, and 70+ dB. For our analyses, these categories were converted to noise levels based on their midpoints (and 55 dB and 70 dB for the lowest and highest ends of the range, respectively).

The survey included 13 annoyance questions, all introduced by the prompt: *“Thinking about the last 12 months or so, when you are here at home, how much does each of the following bother, disturb or annoy you?”.* In addition to the primary item “*Noise from aircraft*”, annoyance from three other noise sources was assessed: “*Noise from cars, trucks or other road traffic*”, “*Your neighbors’ noise or other activities*”, and “*Any other noises you hear when you are here at home*”. Additional items explored non-noise annoyances: “*Smells or dirt from road traffic*”, “*Smoke, gas or bad smells from anything else*”, “*Litter or poorly kept up housing*”, “*Undesirable business, institutional or industrial property*”, “*A lack of parks or green spaces*”, “*Inadequate public transportation*”, “*The amount of neighborhood crime*”, “*Poor city or county services*”, and “*Any other problems that you notice when you are here at home*”. For each item, the possible responses were rated on a 5-point scale: “*Not at all*”, “*Slightly*”, “*Moderately*”, “*Very*” and “*Extremely*”. For our analysis of the aviation noise annoyance item, we followed the NES approach and grouped “*Very*” and “*Extremely*” response levels into a single “*High annoyance*” category, classifying all other response as “*Not highly annoyed*”. We also calculated an “Annoyance from other noise sources” score. This was computed by summing the 5-point scale (1= Not at all annoyed to 5=Extremely annoyed) responses for the three non-aviation noise items (roadway, neighbors, and other noises), yielding a possible score range of 3 to 15 (5 points per item). Additionally, we computed a “non-noise annoyance” score by applying the same 5-point approach and summing the responses for the non-noise annoyance items listed previously. Finally, a cumulative “total annoyance” score was computed by summing the 5-point scale responses for all 13 items.

In addition to the annoyance questions, the NES asked survey participants to provide an overall rating of their neighborhood using this question: “*Now considering how you feel about everything in your neighborhood, how would you rate your neighborhood as a place to live on a scale from 0 to 10 where 0 is worst and 10 is best?*” The questionnaire form clearly indicated that 0 represented “*Worst*” and 10 represented “*Best*”. For our analysis, we considered the statistical distribution of responses, which indicated a median response of 7, and a first quartile of 6. Given that, on the questionnaire form, the response option of 5 was presented as a middle rating, we considered ratings less than 5 to signify a “worse” rating of their neighborhood.

Our statistical approach included a Principal Components Analysis (PCA) of the 5-point scale responses to the annoyance items to identify key features explaining variation in the responses. PCA loadings were used to assess the feasibility of aggregating the questionnaire items into aviation noise annoyance, and scores for annoyance from other noise sources versus non-noise annoyance. To confirm the dose-response curve for high annoyance from aircraft noise, we conducted a logistic regression analysis of the binary outcome (high annoyance from aviation noise) as explained by the noise exposure variable (DNL). We use the term “dose-response curve” to maintain consistency with NES terminology; however, these relationships are often referred to as exposure-response functions in the international noise literature.

Statistical significance was defined as p-values <0.05. Effect measures are reported along with 95% confidence intervals, calculated from standard errors. After fitting the main dose-response curve, secondary analyses were conducted. These included assessing: (a) an interaction between the DNL variable and the score for annoyance from other noise sources, and (b) an interaction between the DNL variable and the score for non-noise annoyance. These interactions were assessed to determine possible modification of the dose-response curve to other types of annoyances.

Our statistical approach for the neighborhood rating outcome employed logistic regression analysis. We conducted logistic regression of the binary outcome (“worse” neighborhood rating, defined as a rating <5) as explained by noise exposure variable (DNL). We also conducted secondary analyses considering other explanatory variables, including (a) aviation noise annoyance on a 5-point scale and (b) the cumulative total annoyance score.

All analyses were conducted in R version 4.5.0.

## Results and Discussion

Of the 10,007 survey responses, 7,278 (72.7%) provided complete responses to all annoyance items and had estimated noise exposures (Table 1). Among these, 30 did not have responses to the neighborhood rating. Of the various annoyance items, the greatest source of high annoyance was attributed to noise from aircraft (N=2,805, 38.5%). Other sources of noise contributed to some, but not as much high annoyance (noise from road traffic N=1,060, 14.6%; noise from neighbors N=887, 12.2%; and other noise N=1,120, 15.5%). Among non-noise annoyance items, the most prevalent causes of high annoyance were litter (N=1,433, 19.7%), other problems (N=1,385, 19.0%), and crime (N=1,299, 17.8%).

**Table 1.** Summary of the annoyance items, cumulative annoyance scores, and neighborhood rating by Aviation DNL noise exposure categories.

| DNL Noise Exposure |  |  |  |  |  |  |
| --- | --- | --- | --- | --- | --- | --- |
|  | ≤55 dB<br>(N=2578) | 55 - 60 dB<br>(N=2405) | 60 - 65 dB<br>(N=1455) | 65 - 70 dB<br>(N=629) | 70+ dB<br>(N=211) | Overall<br>(N=7278) |
| Noise from aircraft |  |  |  |  |  |  |
| Not highly annoyed | 1988 (77.1%) | 1482 (61.6%) | 701 (48.2%) | 242 (38.5%) | 60 (28.4%) | 4473 (61.5%) |
| Highly annoyed | 590 (22.9%) | 923 (38.4%) | 754 (51.8%) | 387 (61.5%) | 151 (71.6%) | 2805 (38.5%) |
| Noise from road traffic |  |  |  |  |  |  |
| Not highly annoyed | 2236 (86.7%) | 2118 (88.1%) | 1187 (81.6%) | 521 (82.8%) | 156 (73.9%) | 6218 (85.4%) |
| Highly annoyed | 342 (13.3%) | 287 (11.9%) | 268 (18.4%) | 108 (17.2%) | 55 (26.1%) | 1060 (14.6%) |
| Noise from neighbors |  |  |  |  |  |  |
| Not highly annoyed | 2287 (88.7%) | 2130 (88.6%) | 1263 (86.8%) | 542 (86.2%) | 169 (80.1%) | 6391 (87.8%) |
| Highly annoyed | 291 (11.3%) | 275 (11.4%) | 192 (13.2%) | 87 (13.8%) | 42 (19.9%) | 887 (12.2%) |
| Other noise |  |  |  |  |  |  |
| Not highly annoyed | 2205 (85.5%) | 2036 (84.7%) | 1221 (83.9%) | 522 (83.0%) | 174 (82.5%) | 6158 (84.6%) |
| Highly annoyed | 373 (14.5%) | 369 (15.3%) | 234 (16.1%) | 107 (17.0%) | 37 (17.5%) | 1120 (15.4%) |
| Smells from traffic |  |  |  |  |  |  |
| Not highly annoyed | 2375 (92.1%) | 2242 (93.2%) | 1304 (89.6%) | 562 (89.3%) | 172 (81.5%) | 6655 (91.4%) |
| Highly annoyed | 203 (7.9%) | 163 (6.8%) | 151 (10.4%) | 67 (10.7%) | 39 (18.5%) | 623 (8.6%) |
| Other smells |  |  |  |  |  |  |
| Not highly annoyed | 2298 (89.1%) | 2164 (90.0%) | 1267 (87.1%) | 536 (85.2%) | 150 (71.1%) | 6415 (88.1%) |
| Highly annoyed | 280 (10.9%) | 241 (10.0%) | 188 (12.9%) | 93 (14.8%) | 61 (28.9%) | 863 (11.9%) |
| Litter |  |  |  |  |  |  |
| Not highly annoyed | 2085 (80.9%) | 1949 (81.0%) | 1159 (79.7%) | 494 (78.5%) | 158 (74.9%) | 5845 (80.3%) |
| Highly annoyed | 493 (19.1%) | 456 (19.0%) | 296 (20.3%) | 135 (21.5%) | 53 (25.1%) | 1433 (19.7%) |
| Undesirable business |  |  |  |  |  |  |
| Not highly annoyed | 2398 (93.0%) | 2218 (92.2%) | 1325 (91.1%) | 572 (90.9%) | 187 (88.6%) | 6700 (92.1%) |
| Highly annoyed | 180 (7.0%) | 187 (7.8%) | 130 (8.9%) | 57 (9.1%) | 24 (11.4%) | 578 (7.9%) |
| Lack of parks |  |  |  |  |  |  |
| Not highly annoyed | 2220 (86.1%) | 2064 (85.8%) | 1208 (83.0%) | 506 (80.4%) | 159 (75.4%) | 6157 (84.6%) |
| Highly annoyed | 358 (13.9%) | 341 (14.2%) | 247 (17.0%) | 123 (19.6%) | 52 (24.6%) | 1121 (15.4%) |
| Inadequate public transit |  |  |  |  |  |  |
| Not highly annoyed | 2243 (87.0%) | 2071 (86.1%) | 1244 (85.5%) | 542 (86.2%) | 174 (82.5%) | 6274 (86.2%) |
| Highly annoyed | 335 (13.0%) | 334 (13.9%) | 211 (14.5%) | 87 (13.8%) | 37 (17.5%) | 1004 (13.8%) |
| Crime |  |  |  |  |  |  |
| Not highly annoyed | 2138 (82.9%) | 2019 (84.0%) | 1166 (80.1%) | 499 (79.3%) | 157 (74.4%) | 5979 (82.2%) |
| Highly annoyed | 440 (17.1%) | 386 (16.0%) | 289 (19.9%) | 130 (20.7%) | 54 (25.6%) | 1299 (17.8%) |
| Poor city/county services |  |  |  |  |  |  |
| Not highly annoyed | 2187 (84.8%) | 2056 (85.5%) | 1212 (83.3%) | 520 (82.7%) | 164 (77.7%) | 6139 (84.4%) |
| Highly annoyed | 391 (15.2%) | 349 (14.5%) | 243 (16.7%) | 109 (17.3%) | 47 (22.3%) | 1139 (15.7%) |
| Other problems |  |  |  |  |  |  |
| Not highly annoyed | 2130 (82.6%) | 1948 (81.0%) | 1159 (79.7%) | 495 (78.7%) | 161 (76.3%) | 5893 (81.0%) |
| Highly annoyed | 448 (17.4%) | 457 (19.0%) | 296 (20.3%) | 134 (21.3%) | 50 (23.7%) | 1385 (19.0%) |

|  | ≤55 dB<br>(N=2578) | 55 - 60 dB<br>(N=2405) | 60 - 65 dB<br>(N=1455) | 65 - 70 dB<br>(N=629) | 70+ dB<br>(N=211) | Overall<br>(N=7278) |
| --- | --- | --- | --- | --- | --- | --- |
| **Annoyance from other (non-aviation) noise sources, score** |  |  |  |  |  |  |
| Mean (SD) | 6.10 (2.79) | 6.11 (2.75) | 6.41 (2.96) | 6.47 (3.00) | 7.09 (3.26) | 6.23 (2.85) |
| Median [Min, Max] | 5.00 [3.00, 15.0] | 6.00 [3.00, 15.0] | 6.00 [3.00, 15.0] | 6.00 [3.00, 15.0] | 7.00 [3.00, 15.0] | 6.00 [3.00, 15.0] |
| **Non-noise annoyance, score** |  |  |  |  |  |  |
| Mean (SD) | 17.2 (7.21) | 17.3 (7.24) | 18.2 (7.77) | 18.6 (7.87) | 20.4 (8.67) | 17.6 (7.47) |
| Median [Min, Max] | 15.0 [9.00, 45.0] | 15.0 [9.00, 45.0] | 17.0 [9.00, 45.0] | 17.0 [9.00, 45.0] | 18.0 [9.00, 45.0] | 16.0 [9.00, 45.0] |
| **Total annoyance, score** |  |  |  |  |  |  |
| Mean (SD) | 25.9 (9.83) | 26.5 (9.86) | 28.1 (10.6) | 28.9 (10.6) | 31.6 (11.5) | 26.9 (10.2) |
| Median [Min, Max] | 24.0 [13.0, 65.0] | 24.0 [13.0, 65.0] | 26.0 [13.0, 65.0] | 27.0 [13.0, 65.0] | 29.0 [13.0, 65.0] | 25.0 [13.0, 65.0] |
| **Neighborhood Rating** |  |  |  |  |  |  |
| Not worse | 2311 (89.6%) | 2117 (88.0%) | 1265 (86.9%) | 544 (86.5%) | 169 (80.1%) | 6406 (88.0%) |
| Worse | 260 (10.1%) | 277 (11.5%) | 184 (12.6%) | 80 (12.7%) | 41 (19.4%) | 842 (11.6%) |
| Missing | 7 (0.3%) | 11 (0.5%) | 6 (0.4%) | 5 (0.8%) | 1 (0.5%) | 30 (0.4%) |

The proportion of survey respondents reporting high annoyance tended to increase with increasing aviation DNL noise exposure for some annoyance items (Table 1). Notably, high annoyance to noise from aircraft increased from 22.9% of those exposed to ≤55 dB DNL to 71.6% of those exposed to 70+ dB DNL. Such a striking increase in high annoyance with increasing aviation DNL noise exposure was not observed for noise exposure from road traffic, noise from neighbors, or other noise. Interestingly, some non-noise related annoyance items tended to show proportions of highly annoyed that increased monotonically with aviation DNL noise exposure, including annoyance with undesirable businesses, lack of parks, and other problems. The highest aviation DNL noise exposure category tended to have the highest proportions of high annoyance for all items – not only for annoyance to noise from aircraft – indicating that residents most exposed to the aircraft noise may experience cumulative annoyance.

Table 1 also provides descriptive statistics for three cumulative annoyance scores. The first is an aggregation of the annoyance from other (non-aviation) noise sources. We find that that even though this score does not account for the annoyance to aviation noise item, it still monotonically increases with increasing aviation DNL noise exposure. This indicates that those exposed to more aviation noise tend to be annoyed to other sources of noise, which may be due to various factors, including airport neighborhoods serving as transportation hubs, which bring together multiple modes of travel, creating more roadway and other noise, as well as increased population density that may also contribute to noise.

Another cumulative annoyance score accounts for the aggregation of the non-noise annoyance items (Table 1). Once again and somewhat surprisingly, we observe that this cumulative score also increases with increasing aviation DNL noise exposure even though it does not include the annoyance to aviation noise item. This suggests that residents of airport noise-impacted neighborhoods may be cumulatively impacted by various environmental issues, not solely noise. This is consistent with the previous observation that the highest aviation DNL noise exposure category has the highest proportions of high annoyance for all items. Also consistent with the findings for the other noise annoyance score and non-noise annoyance score, we find that the total annoyance score, which aggregates all 13 annoyance items, increases with aviation DNL noise exposure.

Finally, Table 1 provides information on the proportions of survey respondents reporting a worse neighborhood rating for varying aviation DNL noise exposure categories. We find that while there is a monotonic increase in the proportion reporting a worse neighborhood rating with increasing aviation noise exposure, the proportion at the highest exposure level (19.4% reporting a worse neighborhood at the 70+ DNL exposure category) does not reflect the same magnitude of effect as asking directly about annoyance from aircraft noise (71.6% reporting high annoyance at the 70+ DNL exposure category). We will visualize the difference in these effects in the modeling results later.

Figure 1 illustrates the dose-response relationship for high annoyance to aircraft noise for aviation DNL noise exposure fit using logistic regression to the pooled annoyance data from the 20 airports. The 95% confidence intervals are narrower than those estimated in the initial NES analyses, which considered inter-airport random effects. Dose-response relationships for the FICON, TNO, and ISO (with 65 dB constant parameter) are also shown in the figure to clearly illustrate the most recent NES-derived curve estimates a much higher percentage of highly annoyed across the 50 – 75 dB DNL range.

**Figure 1.**
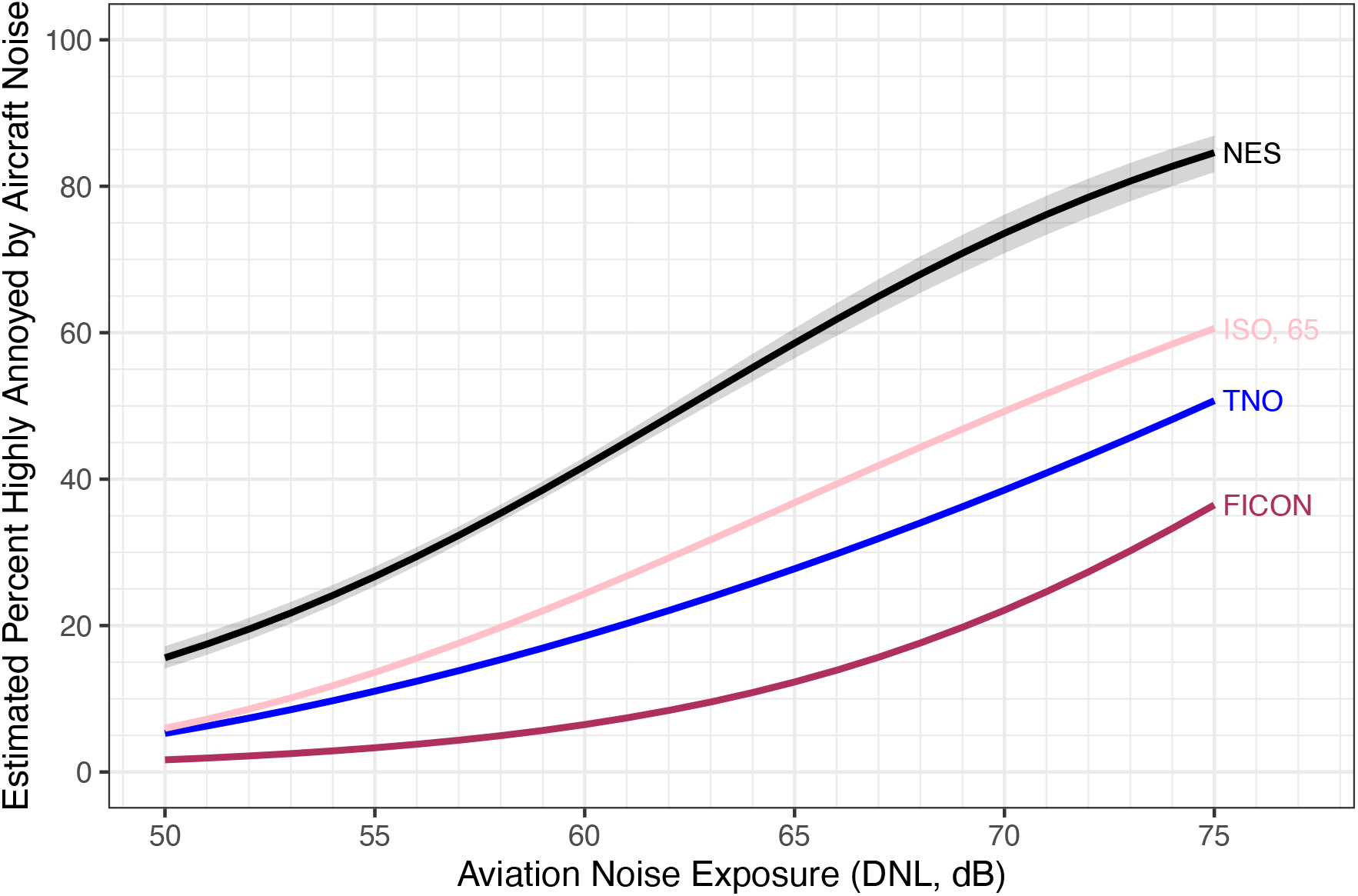
Dose-response for percent highly annoyed by aircraft noise for aviation DNL noise exposure with gray region indicating the 95% CI estimated from a pooled, not airport-specific model. Dose-response relationships for FICON, TNO, and ISO (with 65 dB constant parameter) are also shown for comparison. Model coefficients provided in Table S1.

**Figure 2.**
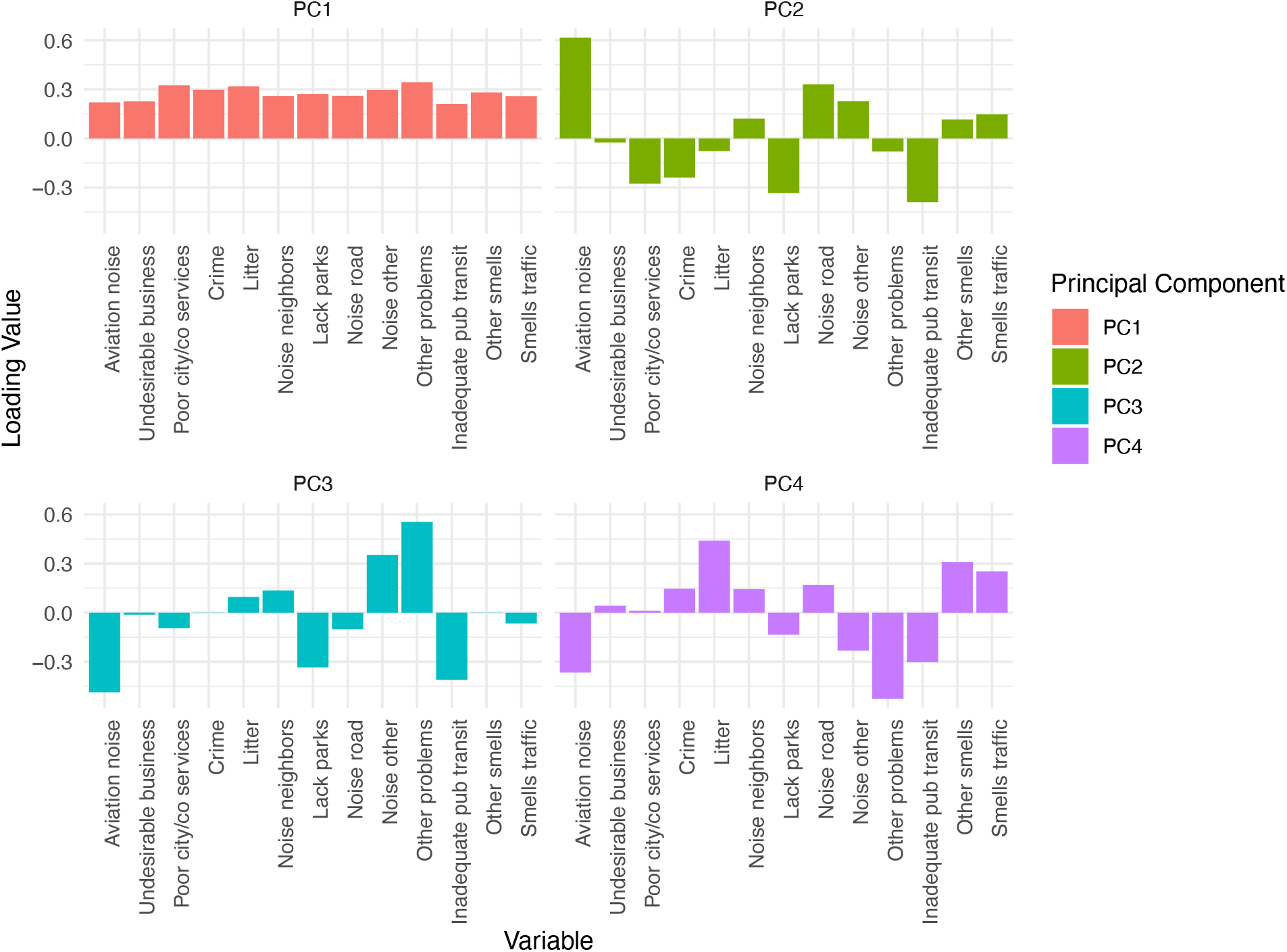
Loadings of variables for annoyance items for the first four PCA components.

PCA analyses of the annoyance items (using 5-point scales) revealed that the first component (PC1) explained over 40% of the variation in the original responses, with the remaining components each explaining <10% of the variation (scree plot in Figure S1). Loadings for the first component (PC1 in Figure 1) were consistent with the presence of cumulative annoyance, with positive loadings observed across all variables. In contrast, the second component (PC2) tended to load heavily on aviation noise annoyance. Components PC3 and PC4 tended to load mostly on non-noise community issues, such as on other problems in the case of PC3, and on litter for PC4.

Given the consistent evidence from PCA and the descriptive statistics that cumulative annoyance is being reported in the NES responses, we re-ran the dose-response logistic regression models with interaction on various cumulative annoyance scores. Figure 3 illustrates the effect of annoyance from other noise sources on the aviation dose-response relationship. Although there was a tendency for those with higher annoyance to other noise sources to have a higher annoyance response to aviation noise (i.e. sensitization), the interaction term of the model was not statistically significant (Table S2). Figure 4 illustrates a similar effect when the cumulative annoyance score from non-noise items is used as the interaction term; those that are more annoyed by non-noise issues tend to be sensitized and have higher annoyance response to aviation noise.

**Figure 3.**
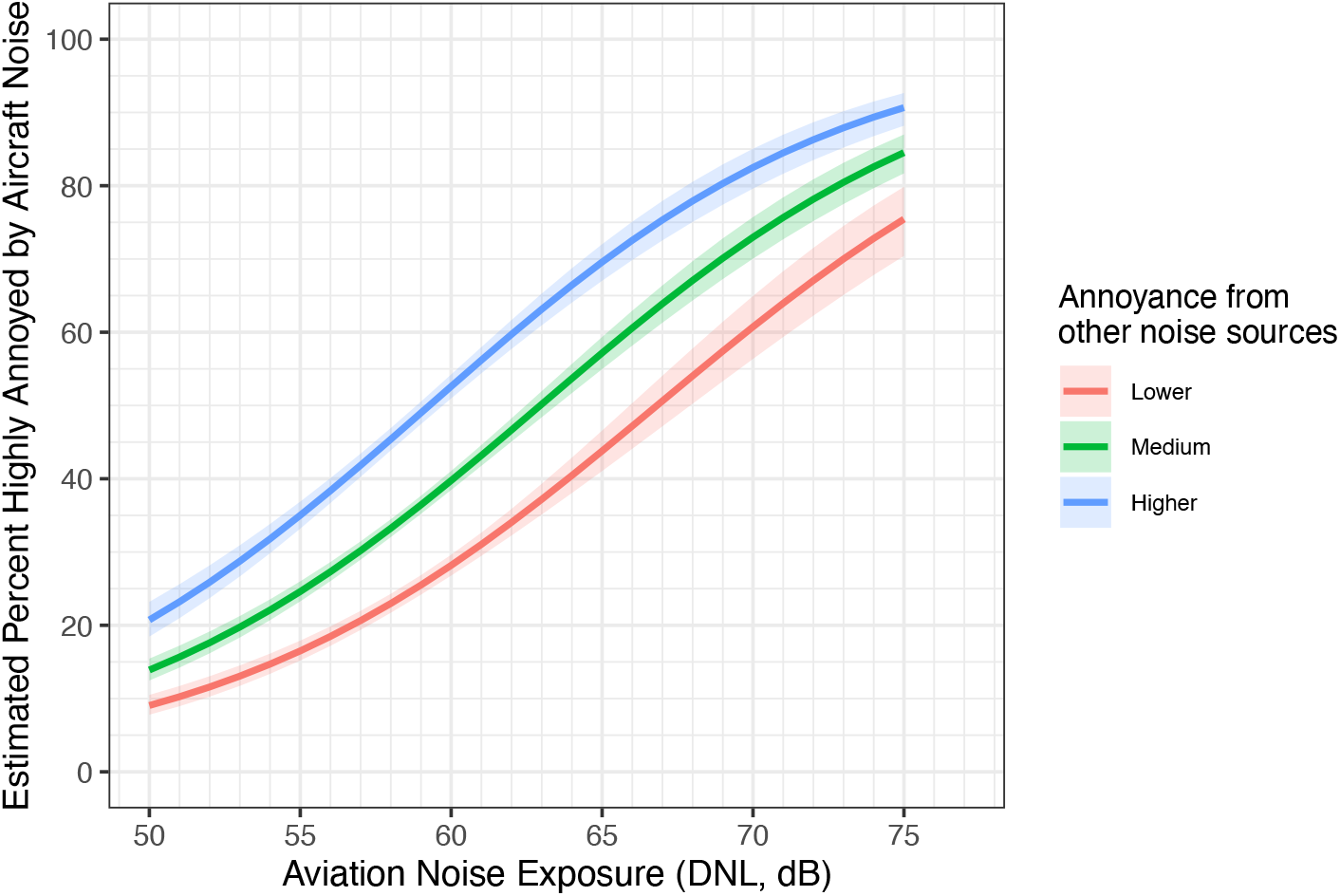
Dose-response for percent highly annoyed by aircraft noise for aviation DNL noise exposure, for those reporting low, medium, or high annoyance to other noise sources (e.g., roadway traffic, neighbors, or other noise). Levels for lower, medium, and higher were based on 25^th^, 50^th^, and 75^th^ percentiles of the other (non-aviation) noise sources annoyance score. Shaded regions indicate the 95% CI estimated from a pooled, not airport-specific model. Model coefficients provided in Table S2.

**Figure 4.**
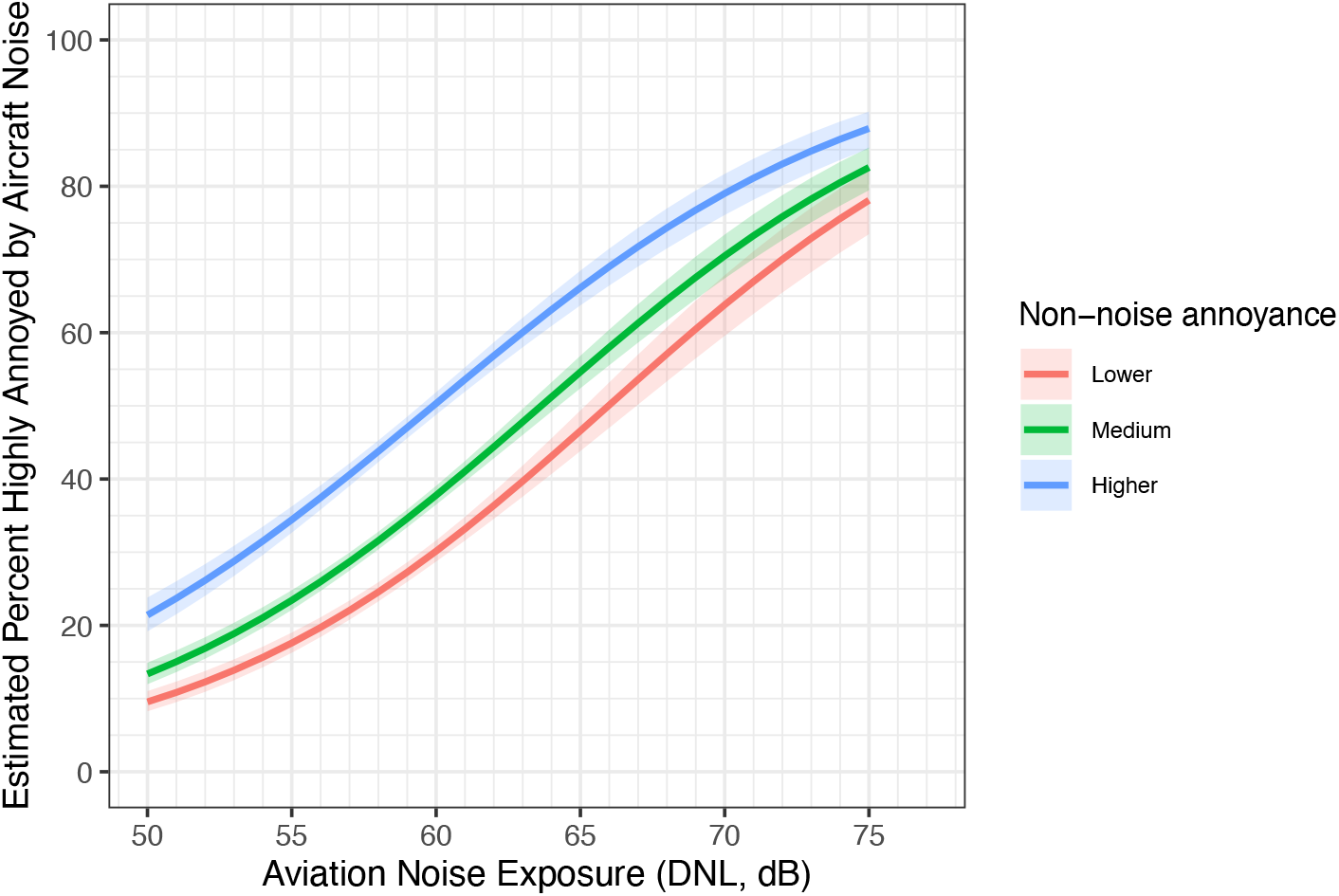
Dose-response for percent highly annoyed by aircraft noise for aviation DNL noise exposure, for those reporting low, medium, or high annoyance to non-noise items (e.g., litter, crime, city services, etc.). Levels for lower, medium, and higher were based on 25^th^, 50^th^, and 75^th^ percentiles of the non-noise annoyance score. Shaded regions indicate the 95% CI estimated from a pooled, not airport-specific model. Model coefficients provided in Table S3.

Similar logistic regression models were explored using the neighborhood rating of “worse” (i.e., ratings <5) as an outcome. Although statistically significant, aviation DNL noise exposure did not explain much of the variation in worse neighborhood ratings, even when considering interaction with other (non-aviation) noise sources (Figure 5). Similarly, there was lack of strong explanation when annoyance to aviation noise was used instead of aviation DNL noise exposure (Figure S2). However, a strong dose-response effect was observed when cumulative total annoyance was considered (Figure 6). These findings indicate that while exposure to aviation noise is highly predictive of aircraft noise annoyance, neither aviation noise exposure nor aircraft noise annoyance alone, are as important as cumulative total annoyance in explaining overall neighborhood ratings.

**Figure 5.**
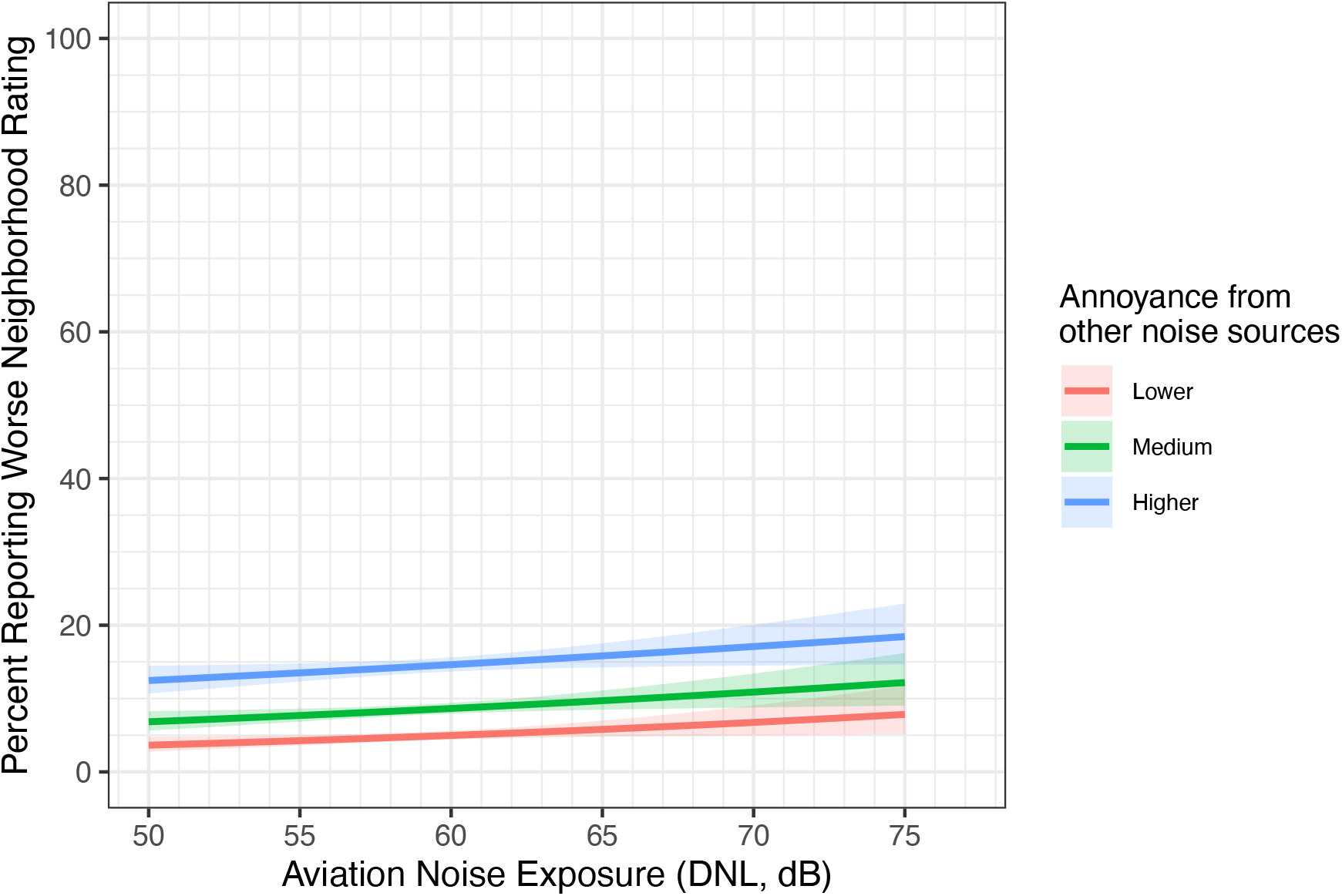
Dose-response for percent reporting a worse neighborhood rating (rating <5) for aviation DNL noise exposure, for those reporting low, medium, or high annoyance to other noise sources (e.g., roadway traffic, neighbors, or other noise). Levels for lower, medium, and higher were based on 25^th^, 50^th^, and 75^th^ percentiles of the other (non-aviation) noise sources annoyance score. Shaded regions indicate the 95% CI estimated from a pooled, not airport-specific model. Model coefficients provided in Table S4.

**Figure 6.**
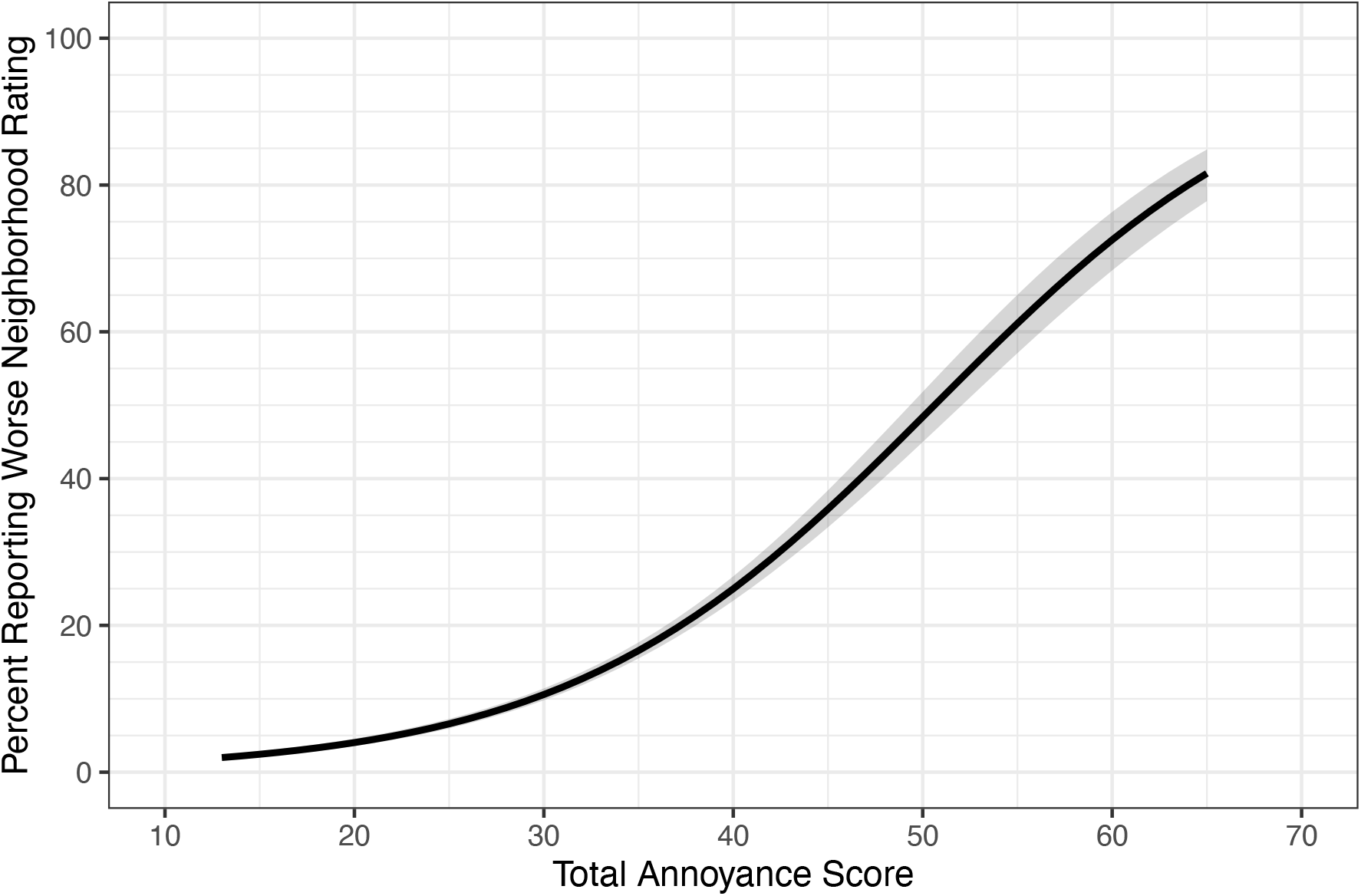
Dose-response for percent reporting a worse neighborhood rating (rating <5) for cumulative total annoyance score. Shaded region indicates the 95% CI estimated from a pooled, not airport-specific model. Model coefficients provided in Table S6.

Our findings shed new light on the potential synergistic relationship among various noise exposures, cumulative annoyance from both noise and non-noise issues, and their impact on population health. While our findings suggest that cumulative noise and cumulative annoyance warrant further attention, the concept of cumulative noise exposure itself is not new. Rice (1977) wrote on the prediction of annoyance from aircraft and traffic noise (12). This experimental study addressed the question of whether a unified dose-response might be appropriate without considering of noise source. That study and subsequent research have found clear evidence for distinct dose-response relationships for high annoyance for different noise sources. For example, Miedema and Oudshoorn (7) examined annoyance from transportation noise and observed separate, approximately polynomial-shaped relationships between DNL (and the daytime, evening, and nighttime noise metric (DENL)) for air, road, and rail noise exposure based on a compilation of noise exposure and annoyance studies. While the focus of their work was on fitting dose-response curves separately for each noise source, they also discussed the importance of equity and policy, noting that equity may emphasize equal tolerance to effects and to specific/local circumstances. They also noted that elaboration of dose-response curve-fitting could include other exposure variables. While our study did not directly include other noise exposures but rather relied on annoyance from road traffic, neighbors, and other noise sources as proxies, our findings are consistent with their concern that the high annoyance dose-response curves may be modified by circumstance and other exposure factors.

A systematic review on noise and annoyance conducted for the World Health Organization (WHO) European noise guidelines largely focused on evidence of annoyance from particular noise sources. However, an attempt was made to consider both factors that may modify the impact of noise exposure on annoyance, and the effect of combined noise on annoyance (6). With respect to the former, for aviation noise, only airport changes (e.g., building a new runway, abrupt change in flight operations) were described as potentially important factors to consider in annoyance studies. The review identified only four publications that addressed annoyance from combined noise sources, an insufficient number for meta-analysis. This led the reviewers to suggest considering the dominant noise source when evaluating annoyance. The “dominant source” annoyance framework is supported by Champelovier et al. (13), who showed that the most annoying source emerges when there is a 5 dB difference between the sources. Wothge et al. (14) also found in their study, which considered combined noise from aircraft, road traffic, and rail, that total noise annoyance was largely determined by the dominant and more annoying source. Yet, Lercher (15) argued that the relationship between multiple combined noise sources and annoyance can be complicated: while masking and frame-of-reference problems can lead people to report lower annoyance with combined noise than expected, there can also be higher noise annoyance due to coincident exposures (e.g., to vibration, air pollution, and tone and impulse components of different noise sources).

Over the last decade, international work on noise annoyance has focused on several important developments, including the standardization of annoyance measurement in social surveys and the selection/transfer of relationships between various noise metrics (16). In fact, the wording of the NES annoyance questions aligned with the ICBEN, using the phrasing “*bothered, annoyed, or disturbed*” in the last 12 months. Furthermore, the survey employed a 5-point scale and “*Very*” and “*Extremely*” response categories were scored as “Highly annoyed”, consistent with ICBEN guidelines. Therefore, findings from the NES should harmonize with international efforts to characterize aviation noise annoyance.

ICBEN has recently contributed to the consideration of potential shifts in the annoyance exposure-response curves due to non-acoustic factors. Standardized approaches to assess and harmonize these potential effects are actively under development under ISO TC43/SC1/WG68 (17). Non-acoustic factors typically include attitudes, demographic factors, and other environmental factors that operate at personal, social, and situational/contextual levels (18). For example, Guski (19) describes noise annoyance as a long-term negative evaluation of living conditions with respect noise that is influenced by personal factors such as sensitivity to noise, fear of harm from the noise source, and ability to cope with noise. Social factors, including distrust of noise agencies or authorities, noise history, and residents’ expectations, can also moderate the impact of noise on annoyance. Moreover, Guski (19) mentions that annoyance is partly related to both acoustic and non-acoustic features of the residents’ personal and social situations. Within this conceptual framework, Guski also notes that broad and diverse impacts – associated with concern, bother, displeasure, irritation, discomfort, distress, exasperation, and other negative connotations – can be self-rated and assessed as multiple variables that are summed. This is essentially the approach we have taken with the NES annoyance items by summing various annoyance item responses into cumulative scores (i.e., a noise-related score, a non-noise score, and a total score). Guski (19) further describes how previous work by McKennell (20) on aviation noise annoyance revealed that individuals who self-reported greater sensitivity to noise tended to show significantly higher annoyance than non-sensitive persons for a given level of noise exposure. Our aggregation of the noise-related annoyance items in the NES reflects a multivariate assessment of noise sensitivity, similar to sensitivity scale by Weinstein (21) that uses multiple Likert-format questions (e.g., about noise-related awareness, annoyance, bother, disturbance) to assess sensitivity. With this approach, we observed evidence of a potential shift in the annoyance exposure-response curve, which suggests that those experiencing greater cumulative annoyance to other noise and non-acoustic factors have greater sensitivity to aviation noise. Continued research into non-acoustic factors is needed, especially as others have noted their relationship to equity and environmental justice concerns for noise policy (22).

The International Civil Aviation Organization (ICAO) has also considered non-acoustic factors in aviation-related annoyance (23). While they recognize ICBEN’s work and the standardization of high annoyance assessment in social surveys, ICAO also highlights the value of more elaborate psychometric approaches to obtain a multifaceted perspective on annoyance through multiple subdimensions. The NES annoyance items do not directly address this need, as there is only a single item related to aviation noise annoyance, while the other items address annoyance related to other noises and the non-noise environment. However, the NES annoyance items do help characterize some situation-specific acoustic and non-acoustic factors that the ICAO suggests should be considered.

The overall neighborhood rating is not a commonly discussed metric in noise literature, and its mention is not prominent in WHO, ICBEN, or ICAO work. However, cumulative neighborhood ratings and scores are common in environmental and social sciences research. Examples include the Residential Environment Assessment Tool (REAT), used to assess neighborhood quality and attachment (24); the Neighborhood Environmental Walkability Survey (NEWS), which asks questions related to physical activity (25–27); and the Built Environment Assessment Tool (BEAT) which assesses multiple aspects of the built environment that affect health (28). These survey-based instruments differ from composite indices and cumulative maps that are based on geospatial (indicator) data, such as the Area Deprivation Index (ADI) (29), the social vulnerability index (SVI) (30), and environmental justice tools, such as the Environmental Protection Agency’s EJSCREEN (31) and California’s CalEnviroScreen (32).

While these instruments rely on multiple questions (and multiple indicator data in the case of mapping tools) to assess different domains, potentially offering improved validity, the NES only included a single question related to overall neighborhood rating. Despite this, we observed in our study that aviation noise exposure and annoyance from aircraft noise, alone, did not strongly influence neighborhood rating. Instead, we found that the overall neighborhood rating was more influenced by the cumulative total annoyance score.

Our study had some limitations. Principally, it utilized de-identified data from the NES public-use file, which provided only categorized noise exposures in 5 dB increments and lacked geolocation coordinates or airport identifiers for individual survey responses to protect personal identification. Despite this limitation, we successfully developed an annoyance dose-response relationship based on the pooled data that exhibits characteristics similar to the FAA’s dose-response curve for NES that was based on a mixed effects model. The absence of geolocation coordinates, however, prevented a more direct assessment of combined noise exposure using comprehensive transportation noise models (i.e., modeled exposures to not only aviation noise but also road and rail noise), such as from the National Transportation Noise Exposure Map (33,34). Such an assessment would have facilitated more direct evaluation of effect modification and sensitivity to cumulative noise. Nevertheless, the NES annoyance items for roadway, household, and other noise sources offered a distinct, and arguably better, perspective on sensitization related to perceived noise impact. Finally, as noted previously, the inclusion of additional questions related to various domains of neighborhood quality might have provided further insight and improved validity compared to the single neighborhood rating question in the NES.

Despite these limitations, the current study offers significant strengths and novel contributions: (1) it is one of the few recent US-based noise studies of annoyance to aircraft noise that relies on a comprehensive sampling of households around many airport communities; (2) it employs annoyance assessments based on current international recommendations; (3) it expands upon the FAA’s NES analysis by considering modification of the dose-response curve by other noise and non-noise factors; and (4) it assesses the relationships among aviation noise exposure, aircraft noise annoyance, cumulative total annoyance, and overall neighborhood rating. Furthermore, the findings of this study underscore the continued need for research that examines effect modification of aviation annoyance dose-response relationships in diverse situational contexts. The findings also indicate that more research is needed to assess the role of aviation other noise sources in cumulative annoyance, in relation to other community stressors and their influence on how residents rate their neighborhoods.

## Conclusion

A clear dose-response relationship between aviation DNL noise exposure and high annoyance to aircraft noise was observed in an analysis of pooled data from the NES. Additionally, this study found evidence of potential effect modification of the aviation dose-response curve by annoyance from other non-noise factors. Although neither aviation noise exposure nor aircraft noise annoyance were found to strongly influence neighborhood rating, a cumulative total annoyance score was found to have a dose-response relationship with neighborhood rating.

## Data Availability

All data produced are available online at:
https://www.airporttech.tc.faa.gov/Products/Airport-Safety-Papers-Publications/Airport-Safety-Detail/ArtMID/3682/ArticleID/2845/Analysis-of-NES

## Acknowledgements

We thank the FAA for providing access to data in the public-use file from the NES.

