## Supplemental Information for "Aviation noise, cumulative annoyance and neighborhood ratings: Effect modification of the annoyance dose-response curve from the Neighborhood Environmental Survey"

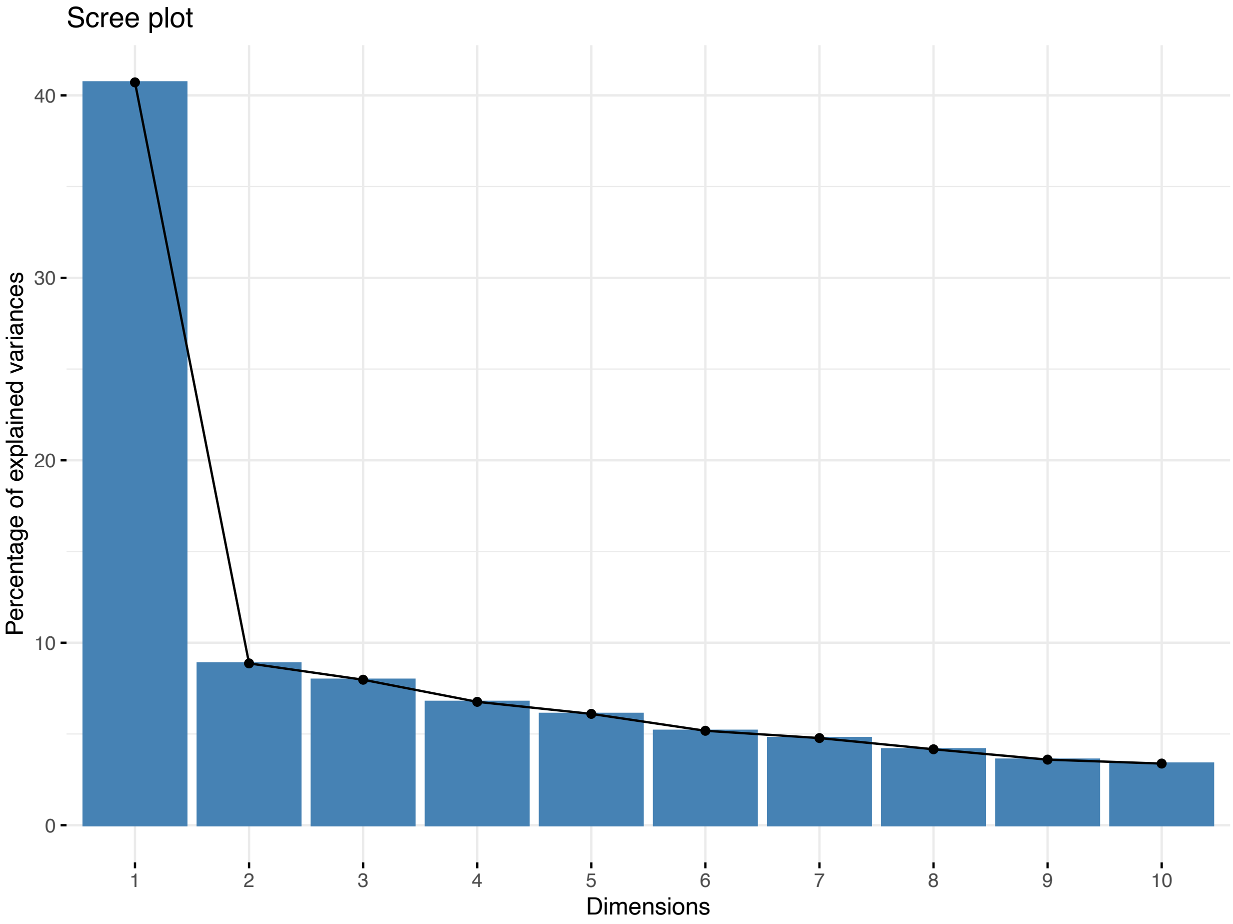


**Figure S1.** Scree plot of PCA of annoyance items.

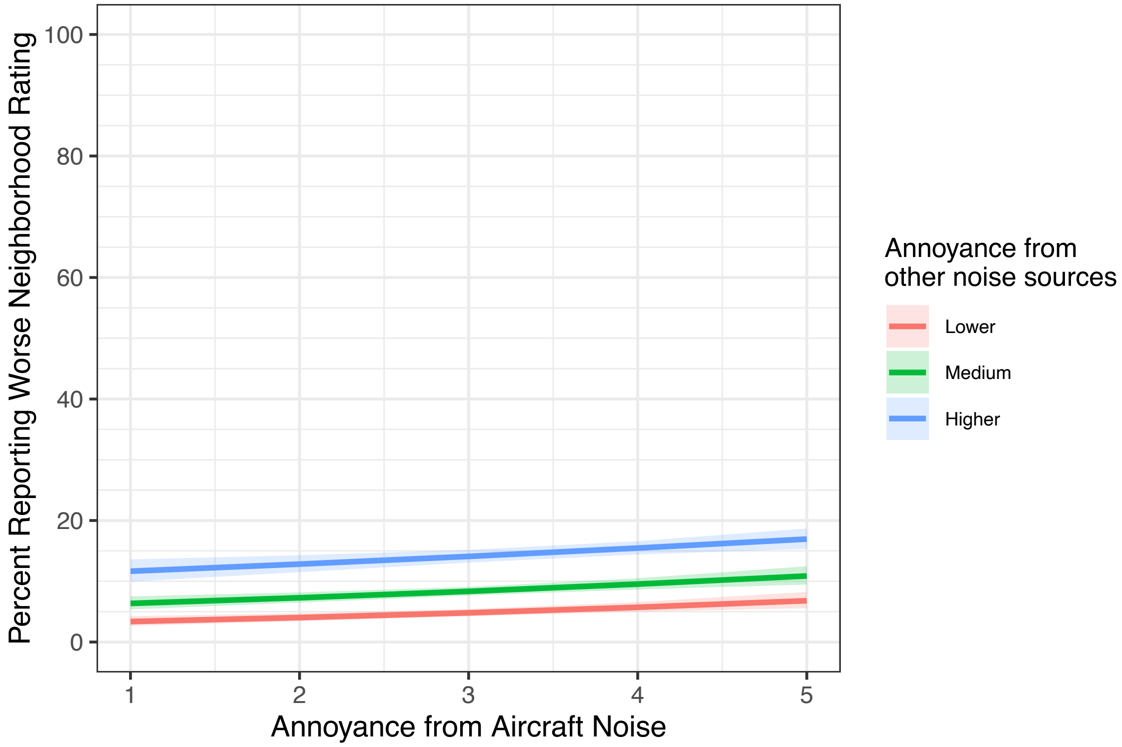


**Figure S2.** Dose-response for percent reporting a worse neighborhood rating (rating <5) for annoyance to aircraft noise score (5-point), for those reporting low, medium, or high annoyance to other noise sources (e.g., roadway traffic, neighbors, or other noise). Levels for lower, medium, and higher were based on 25th, 50th, and 75th percentiles of the other (non-aviation) noise sources annoyance score. Shaded regions indicate the 95% CI estimated from a pooled, not airport-specific model. Model coefficients provided in Table S5.

**Table S1.** Logistic regression model of percentage high annoyance to aircraft noise from aviation DNL noise exposure (dB).

| Characteristic | OR | 95% CI | p-value |
|---|---|---|---|
| (Intercept) | 0.00 | 0.00, 0.00 | <0.001 |
| DNL_continuous | 1.15 | 1.13, 1.16 | <0.001 |

Abbreviations: CI = Confidence Interval, OR = Odds Ratio

**Table S2.** Logistic regression model of percentage high annoyance to aircraft noise from aviation DNL noise exposure (dB) with interaction on cumulative annoyance from other (non-aviation) noises score.

| Characteristic | OR | 95% CI | p-value |
|---|---|---|---|
| (Intercept) | 0.00 | 0.00, 0.00 | <0.001 |
| DNL_continuous | 1.14 | 1.10, 1.17 | <0.001 |
| TotalMA_noise | 1.16 | 0.88, 1.53 | 0.3 |
| DNL_continuous * TotalMA_noise | 1.00 | 1.00, 1.01 | 0.4 |

Abbreviations: CI = Confidence Interval, OR = Odds Ratio

**Table S3.** Logistic regression model of percentage high annoyance to aircraft noise from aviation DNL noise exposure (dB) with interaction on cumulative non-noise annoyance score.

| Characteristic | OR | 95% CI | p-value |
|---|---|---|---|
| (Intercept) | 0.00 | 0.00, 0.00 | <0.001 |
| DNL_continuous | 1.16 | 1.13, 1.20 | <0.001 |
| Non-noise annoyance, score | 1.15 | 1.04, 1.27 | 0.006 |
| DNL_continuous * Non-noise annoyance, score | 1.00 | 1.00, 1.00 | 0.3 |

Abbreviations: CI = Confidence Interval, OR = Odds Ratio

**Table S4.** Logistic regression model of percentage reporting worse neighborhood rating from aviation DNL noise exposure (dB) with interaction on cumulative annoyance from other (non-aviation) noises score.

| Characteristic | OR | 95% CI | p-value |
|---|---|---|---|
| (Intercept) | 0.00 | 0.00, 0.02 | <0.001 |
| DNL_continuous | 1.05 | 1.00, 1.10 | 0.045 |
| TotalMA_noise | 1.66 | 1.22, 2.25 | 0.001 |
| DNL_continuous * TotalMA_noise | 1.00 | 0.99, 1.00 | 0.2 |

Abbreviations: CI = Confidence Interval, OR = Odds Ratio

**Table S5.** Logistic regression model of percentage reporting worse neighborhood rating from annoyance to aircraft (5-point scale) with interaction on cumulative annoyance from other (non-aviation) noises score.

| Characteristic | OR | 95% CI | p-value |
|---|---|---|---|
| (Intercept) | 0.01 | 0.00, 0.01 | <0.001 |
| MALACX.x | 1.30 | 1.11, 1.52 | 0.001 |
| Annoyance from other (non-aviation) noise sources, score | 1.42 | 1.32, 1.53 | <0.001 |
| MALACX.x * Annoyance from other (non-aviation) noise sources, score | 0.98 | 0.96, 1.00 | 0.044 |

Abbreviations: CI = Confidence Interval, OR = Odds Ratio

**Table S6.** Logistic regression model of percentage reporting worse neighborhood rating from cumulative total annoyance score.

| Characteristic | OR | 95% CI | p-value |
|---|---|---|---|
| (Intercept) | 0.01 | 0.00, 0.01 | <0.001 |
| TotalMAL | 1.11 | 1.10, 1.12 | <0.001 |

Abbreviations: CI = Confidence Interval, OR = Odds Ratio
